# Is it safe to lift COVID-19 travel bans? The Newfoundland story

**DOI:** 10.1101/2020.07.16.20155614

**Authors:** Kevin Linka, Proton Rahman, Alain Goriely, Ellen Kuhl

## Abstract

A key strategy to prevent a local outbreak during the COVID-19 pandemic is to restrict incoming travel. Once a region has successfully contained the disease, it becomes critical to decide when and how to reopen the borders. Here we explore the impact of border reopening for the example of Newfoundland and Labrador, a Canadian province that has enjoyed no new cases since late April, 2020. We combine a network epidemiology model with machine learning to infer parameters and predict the COVID-19 dynamics upon partial and total airport reopening, with perfect and imperfect quarantine conditions. Our study suggests that upon full reopening, every other day, a new COVID-19 case would enter the province. Under the current conditions, banning air travel from outside Canada is more efficient in managing the pandemic than fully reopening and quarantining 95% of the incoming population. Our study provides quantitative insights of the efficacy of travel restrictions and can inform political decision making in the controversy of reopening.

*“There is one and only one way to absolutely prevent it and that is by establishing absolute isolation. It is necessary to shut off those who are capable of giving off the virus from those who are capable of being infected, or vice versa*.*” The Lessons Of The Pandemic, Science 1919*.

## 1 Motivation

On July 3, 2020, the Canadian province of Newfoundland and Labrador enjoyed the rather exceptional and enviable position of having the coronavirus pandemic under control with the total number of 261 cases, with 258 recovered, 3 deaths, no active cases for 16 consecutive days, and no new cases for 36 days [2]. On the same day, after a two-months long local travel ban, the *Atlantic Bubble* opened to allow air travel between the four Atlantic Provinces, Newfoundland and Labrador, Nova Scotia, New Brunswick, and Prince Edward Island, with no quarantine requirements for travelers [37]. Under the increasing pressure to fully reopen, health officials and political decision makers now seek to understand the risk of gradual and full reopening under perfect quarantine conditions and quarantine violation [24].

The province of Newfoundland and Labrador is the second smallest Canadian province with a population of 519,716. It has two major geographical divisions, the island of Newfoundland that accounts for 92% of the population and a continental region of Labrador that is home to the remaining 8% [35]. The demographics of its population, the highest rates of obesity and overweight, metabolic disease, and cancer nationally, and an unhealthy lifestyle with the highest rate of cigarette smoking among all provinces set Newfoundland and Labrador apart from the rest of Canada [3]. These factors are critical when developing policies for the management of COVID-19.

The first reported case of COVID-19 in Newfoundland and Labrador was on March 14, 2020 followed by a rapid escalation in the number of cases caused by a super-spreader event at a funeral home [37]. Rapid and well-coordinated implementation of provincial public health measures resulted in excellent viral epidemic control of the first wave and the province has not had a documented case of community transmission since mid-April 2020 [2]. In the absence of any reported COVID-19 cases in the province, the risk of any future outbreaks will originate from travelers. On May 4, 2020 the Chief Medical Officer of the province issued a Special Measures Order stating that the only people allowed to enter the province were residents of Newfoundland and Labrador, asymptomatic workers, and those granted a permit due extenuating circumstances [31]. Since there is a limited number of entry points into Newfoundland, with three quarters of all travelers entering via air [26], passenger air travel is a good metric to assess the impact of travel restrictions on the importation risk in Newfoundland [15].

A relaxation of the travel ban naturally induces anxiety and fear of a new outbreak. From a public health perspective, the major challenges are *(i)* to understand the effect of the travel bubble; *(ii)* to predict the effect of a wider opening to the rest of Canada and the United States; and *(iii)* to estimate the effect of imperfect quarantine assuming that a fraction of travelers would ignore the guidelines for self-isolation. Answering these questions will help to understand the short- and long-term effects of reopening.

To explore whether and when it would be safe to lift the travel ban, we model the dynamics of COVID-19 using a network approach that links a local epidemiological model with air-traffic mobility [21]. Local epidemiological modeling [17] is now a well-accepted approach to follow the dynamics of a homogeneous population during an epidemic [13]. The extra network layers allow us to capture the mobility between different local populations [22]. A unique feature of this approach is that we can dynamically infer the parameters of the epidemiology model using reported case data [18,38] and update it in real time during the progression of the disease [23]. In addition, we can easily extract mobility data from passenger air travel statistic between the different locations [15]. Here use this approch to study three reopening scenarios by gradually adding air traffic from *(i)* the Atlantic Provinces; *(ii)* all of Canada; and *(iii)* all of North America.

## 2 Methods

### 2.1 Epidemiology modeling

We model the local epidemiology of the COVID-19 outbreak using an SEIR model [5, 29, 41, 42] with four compartments, the susceptible, exposed, infectious, and recovered populations, governed by the set of ordinary differential equations,

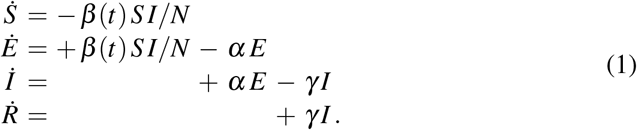

Here 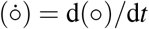 denotes the time-derivative of the compartment 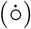 and *N* = *S* + *E* + *I* + *R* is the total population. Three parameters govern the transition from one compartment to the next: the contact rate *β*, the latency rate *α*, and the infectious rate *γ*. They are the inverses of the contact period *B* = 1/*β*, the latent period *A* = 1/*α*, and the infectious *C* = 1/*γ*. For simplicity, we assume that the latency rate *α* = 1/2.5 days^−1^ and the infectious rate *γ* = 6.5 days^−1^ are disease-specific for COVID-19, and constant in space and time [19, 20, 33]. To account for societal and political actions [39], we introduce a behavior specific dynamic contact rate *β* = *β* (*t*) that varies both in space and time [22].

For easier interpretation, we express the contact rate,

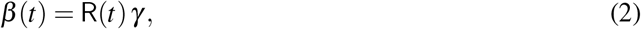

in terms of the dynamic effective reproduction number R(*t*) [9], for which we make an ansatz of Gaussian random walk type [30] with a constant time-window of four days,

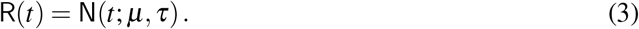

Here N(*t*) is the time-varying Gaussian distribution,

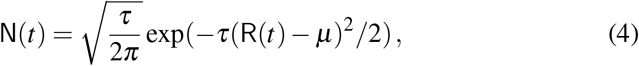

parameterized in terms of the drift *µ* and the daily stepwidth *τ* = *τ*\*/[1.0 −*s*], where *τ*\* is the the step width precision and *s* is the associated smoothing parameter [23].

### 2.2 Mobility modeling

We model each province, territory, and state of North America as a homogeneous population with its own local SEIR dynamics and connect them to the province of Newfoundland and Labrador through a global mobility network [4]. From this mobility network, we create a weighted graph 𝒢 in which the i = 1, ‥, n nodes 𝒩 represent the individual provinces, territories, and states and the weighted edges *ℰ* represent the mobility between them [11]. We approximate the weights of the edges using the average daily passenger air travel statistics [15], and summarize this information in the adjacency matrix *A*_ij_ that reflects the travel frequency between two regions i and j, and in the degree matrix,

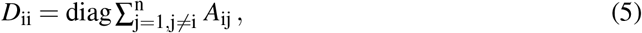

that reflects the number of incoming passengers for each region i. The difference between the degree matrix *D*_ij_ and the adjacency matrix *A*_ij_ defines the weighted graph Laplacian [21],

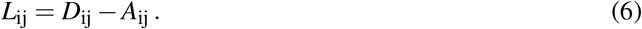

For the province of Newfoundland and Labrador, the incoming and outgoing passenger air travel from and to other regions is relatively similar and we can simply average the two which results in an undirected graph 𝒢 and symmetric adjacency and Laplacian matrices, *A*_ij_ = *A*_ji_ and *L*_ij_ = *L*_ji_. Since we focus on the early phase of a resurgence, we only simulate the mobility into and out of Newfoundland and Labrador and neglect the intraregional mobility between all other regions. This implies that only a single row and column of the adjacency matrix are populated, *A*_i1_ = *A*_1j_ ≠ 0,., *A*_ij_ = 0 for all i, j ≠ 1. Similarly, only one entry of the degree matrix is populated, *D*_11_ ≠ 0, while all other entries are zero, *D*_ii_ = 0 for all i ≠ 1.

Figure 1 illustrates three discrete graphs of the Atlantic Provinces, Canada, and North America. The graph of the Atlantic Provinces 𝒢_AP_ consists of n = 4 nodes and e = 3 edges, shown in dark blue; the graph of Canada 𝒢_CA_ consists of n = 13 nodes and e = 12 edges shown in dark and light blue, and the graph of North America 𝒢_NA_ consists of n = 64 nodes and e = 63 edges shown in dark and light blue and red. We discretize our SEIR model on these weighted graphs 𝒢 and introduce the susceptible, exposed, infectious, and recovered populations *S*_i_, *E*_i_, *I*_i_, and *R*_i_ as global dependent variables at the i = 1, …, n nodes of the graph. This results in a spatial discretization of the set of equations with 4 n unknowns,

**Fig. 1.**
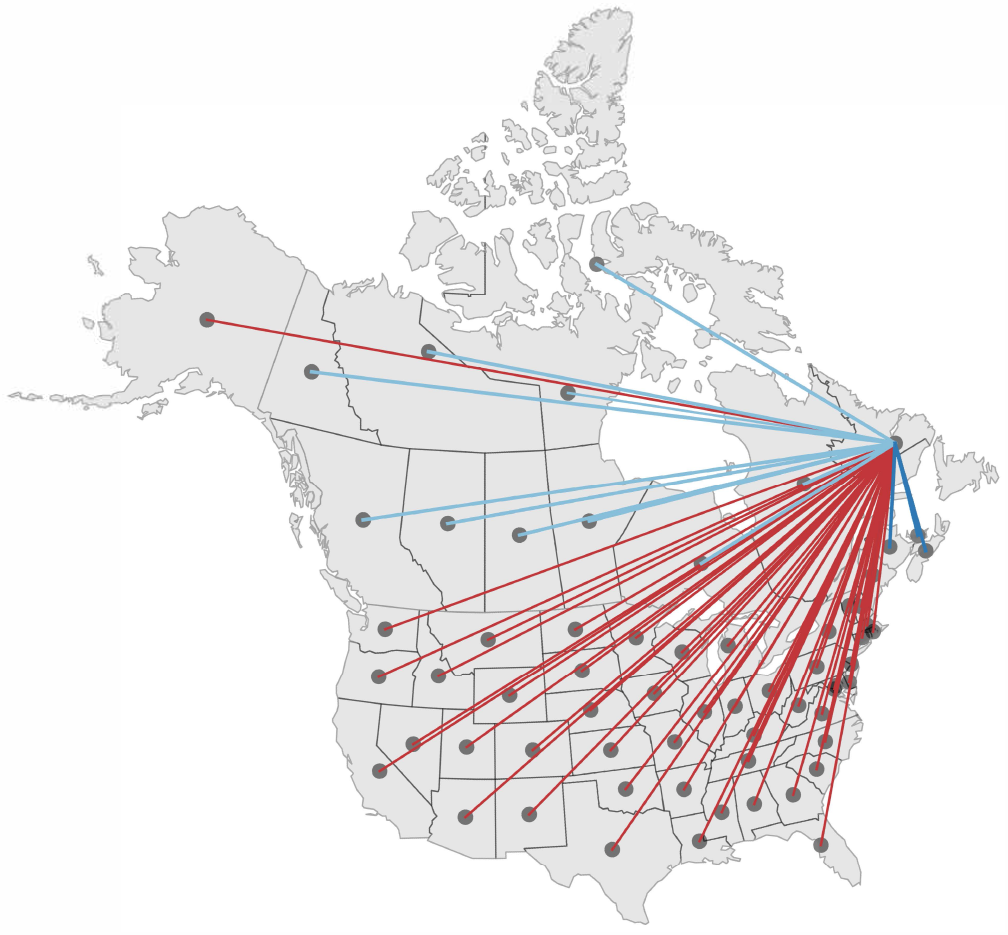
Mobility modeling. Discrete graphs 𝒢_AP_ of the Atlantic Provinces (green), 𝒢_CA_ of Canada (green and red), and 𝒢_NA_ of North America (green, red, and blue) with n = 4, n = 13, and n = 64 nodes and e = n − 1 edges that represent the main travel routes to Newfoundland and Labrador. Dark blue edges represent the connections from the Atlantic Provinces, light blue edges from the other Canadian provinces and territories, and red edges from the United States.

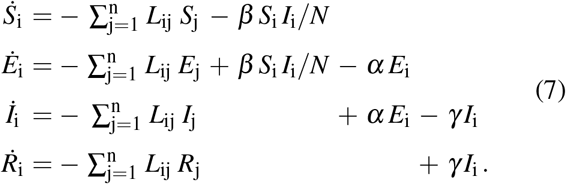

We discretize the system of equations (7) in time using an explicit Euler forward time integration scheme and approximate the time derivatives as 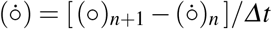, where *Δt* denotes that discrete time step size.

### 2.3 COVID-19 epidemiology and mobility data

For the COVID-19 epidemiology data, we draw the COVID-19 history of all 13 Canadian provinces and territories [2] and all 51 United States [34] from the beginning of the outbreak until July 1, 2020. From these data, we extract the newly confirmed cases Î(*t*) as the difference between today’s and yesterday’s reported cases and the cumulative confirmed cases 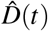 as the sum of all reported cases to date.

For the mobility data, we sample all North American air traffic data to and from the province of Newfoundland and Labrador and use a 15-month period before the COVID-19 outbreak, from January 1, 2019 to March 31, 2020, to estimate the daily air traffic [15]. Figure 2 illustrates the average daily air traffic to and from Newfoundland and Labrador from all of North America. The locations with the largest mobility to and from the province are Ontario with 188/day, Quebec with 146/day, Alberta with 143/day, and Florida with 102/day, followed by New Jersey and British Columbia both with 67/day, Nova Scotia with 41/day, Manitoba and Texas both with 20/day.

**Fig. 2.**
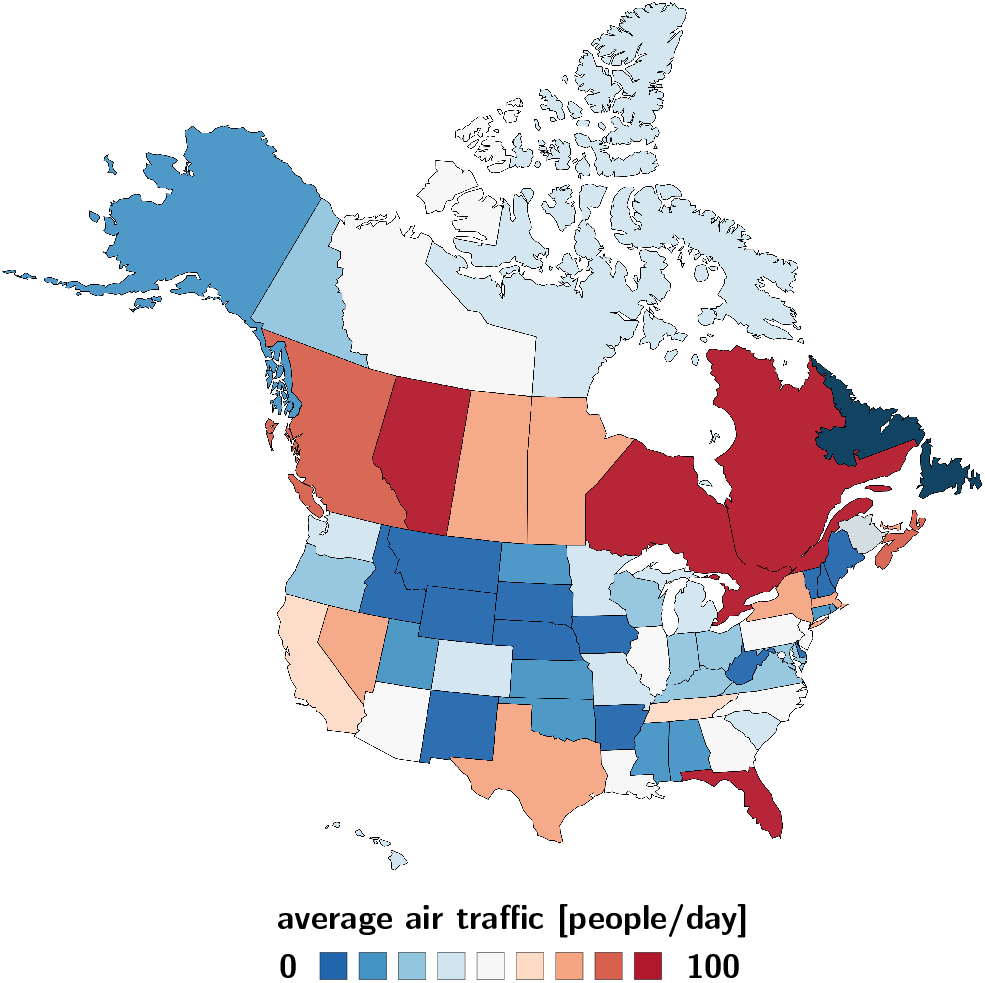
Average daily air traffic to and from Newfoundland and Labrador. Number of daily incoming and outgoing air passengers from the Canadian Provinces and Territories and the United States for a 15-months period before the COVID-19 outbreak, from January 1, 2019 to March 31, 2020, as reported by the International Air Transport Association [15].

### 2.4 Bayesian inference

Our dynamic SEIR model [22] introduces three parameters for each territory, province, or state to characterize the individual time-dependent effective reproduction numbers R(*t*),

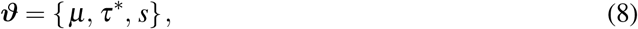

the drift *µ*, the stepwidth precision *τ*^*^, and the smoothing parameter *s* of our Gaussian random walk type ansatz (4). We estimate these model parameters using Bayesian inference [1, 16, 27] with Markov Chain Monte Carlo sampling [21, 29]. Specifically, we adopt a Student’s t-distribution for the likelihood between the reported cumulative case numbers 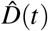 and the simulated cumulative case numbers *D*(*t, ϑ*) = *I*(*t*) + *R*(*t*) with a case-number-dependent width,

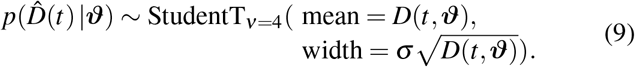

Here *σ* represents the width of the likelihood 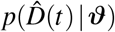 between the reported and simulated case numbers, 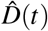 and *D*(*t*, ***ϑ***). We apply Bayes’ rule

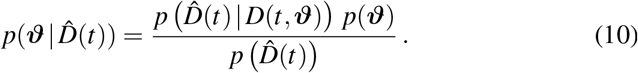

to obtain the posterior distribution of the parameters using the reported case numbers and selected prior distributions using the NO-U-Turn sampler (NUTS) [14] implemented in the Python package PyMC3 [32]. For the prior distributions, we select the drift *µ*∼ Normal(0,2), the stepwidth precision *τ*^*^∼ Exponential (1/2), the smoothing parameter *s* ∼ Uniform(0,1), and the likelihood width *σ*∼ HalfCauchy(*β* = 1). We report the resulting effective reproduction number R(*t*) and the reported cases 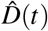 and and simulated cases *D*(*t*) = *I*(*t*)+ *R*(*t*) for each territory, province, and state from the early outbreak on March 15 until July 1, 2020.

### 2.5 Reopening forecast

We explore two reopening scenarios, partial and total airport reopening, with perfect and imperfect quarantine conditions. To model the effect of partial and total reopening, we allow travel within the Atlantic Provinces using the graph 𝒢_AP_, within Canada using the graph 𝒢_CA_, and within North America using the graph 𝒢_NA_ from Figure 1.

To model the effect of perfect and imperfect quarantine, we assume that a fraction *ν*_q_ of all incoming travelers fully satisfies a 14-day quarantine period, whereas [1 − *ν*_q_] violate the quarantine conditions. We modulate the graph Laplacian, 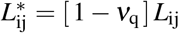, by the fraction of travelers [1− *ν*_q_] that violates the quarantine requirements and select different quarantine levels, *ν*_q_ = [0%,25%, 50%, 75%, 95%]. For *ν*_q_ = 0%, none of the incoming travelers satisfy the quarantine requirements and all incoming exposed and infectious individuals mix instantaneously with the local population, 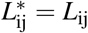. For *ν*_*q*_ = 100%, all incoming travelers satisfy the mandatory quarantine requirements, there is no quarantine violation, and none of the incoming exposed and infectious individuals mix with the local population until they have fully recovered, 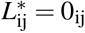. We estimate the effects of both re-opening scenarios on the COVID-19 outbreak dynamics in Newfoundland and Labrador for a 150-day period starting July 1, 2020.

## 3 Results

### Outbreak dynamics of COVID-19 in North America

Figure 3 illustrates the outbreak dynamics of COVID-19 in the Atlantic Provinces, the other Canadian provinces and territories, and the United States from the beginning of the outbreak until July 1, 2020. The top of each graph shows the time-dependent effective reproduction number R(*t*) of our dynamic SEIR model according to equation (4) as red curve,the bottom shows the confirmed cases 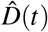 as dots and the model fit *D*(*t*) = *I*(*t*)+ *R*(*t*) according to equations (1.3) and (1.4) as orange curve. The solid lines represent the median values, the shaded areas highlight the 95% the confidence intervals. Table 1 summarizes the outbreak dynamics on July 1, 2020, the susceptible, exposed, infectious, and recovered populations *S, E, I, R* and the effective reproduction number R_t_ for all 64 locations.

**Table 1.**
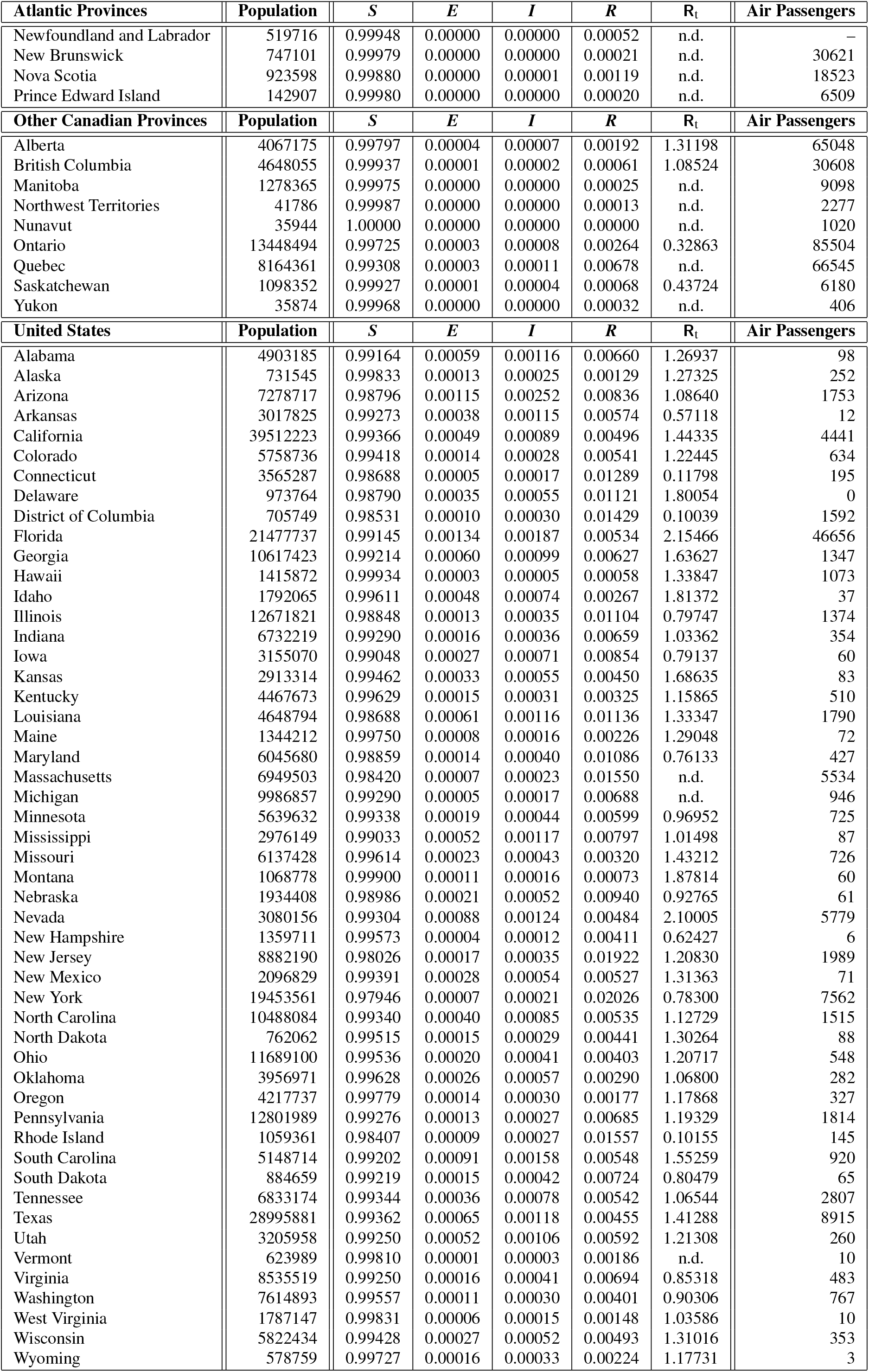
Outbreak dynamics of COVID-19 in the Atlantic Provinces, all other Canadian provinces, and the United States. Susceptible, exposed, infectious, and recovered populations *S, E, I* and *R*, and effective reproduction number R_*t*_ on July 1, 2020 inferred from reported case numbers [2, 34], and air passengers before the COVID-19 outbreak, from January 1, 2019 to March 31, 2020 [15]; n.d. indicates R_*t*_ not defined.

**Fig. 3.**
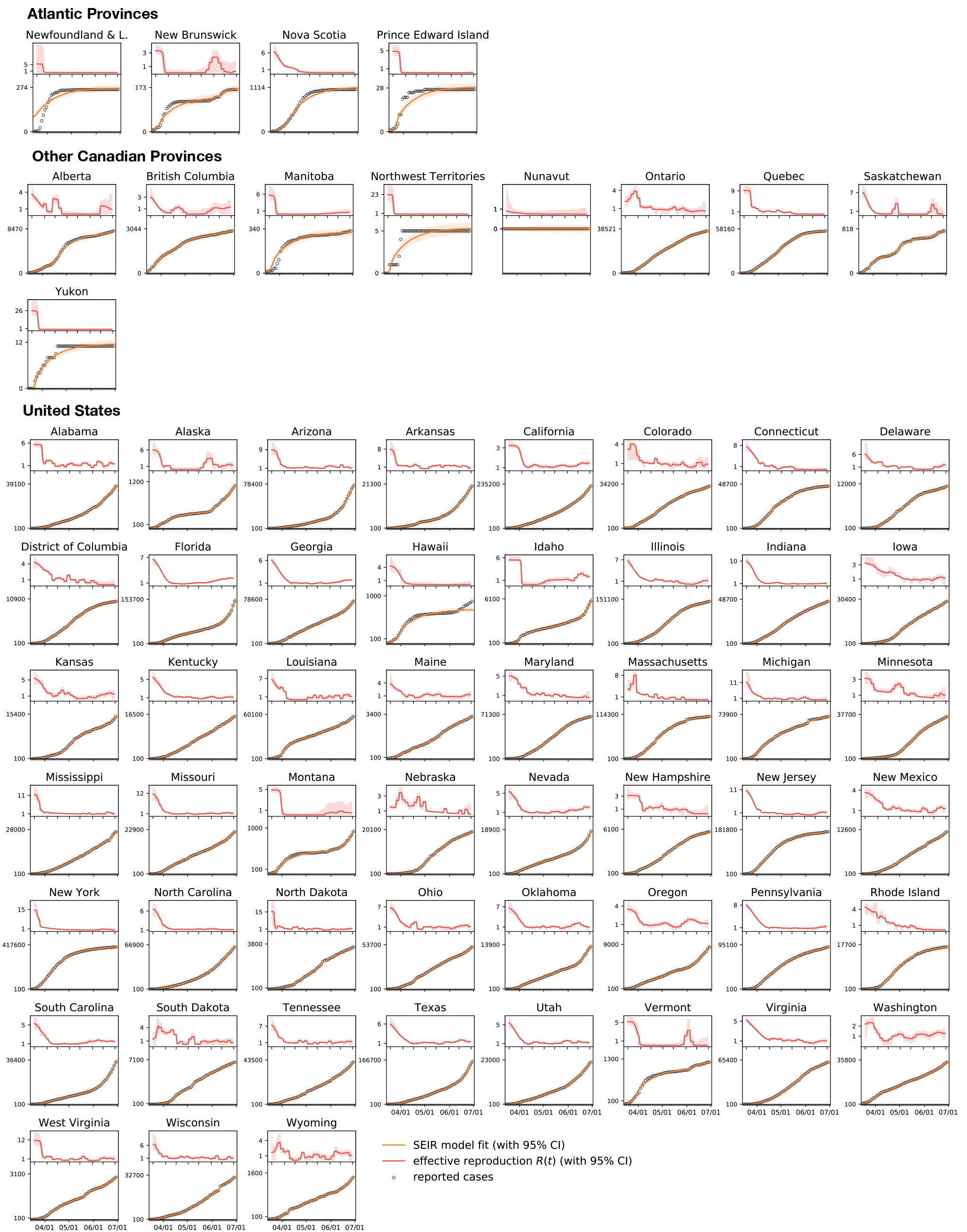
Outbreak dynamics of COVID-19 in the Atlantic Provinces, all other Canadian provinces, and the United States. Effective reproduction number (red), confirmed cases (dots) [2, 34], and model fit (orange) from the beginning of the outbreak until July 1, 2020. Solid lines represent the median values, shaded areas highlight the 95% confidence intervals.

Of most interest are the regional exposed and infectious populations *E* and *I* on the day of reopening, columns 4 and 5 of Table 1, as they determine how rapidly the outbreak will travel into Newfoundland and Labrador. The exposed population on July 1, 2020 was largest in Florida with 0.134%, Arizona with 0.115%, South Carolina with 0.091%, Nevada with 0.088%, and Texas with 0.065%. The infectious population on July 1, 2020 was largest in Arizona with 0.252%, Florida with 0.187%, South Carolina with 0.158%, Nevada with 0.124%, and Texas with 0.118%. For comparison, no Canadian province or territory had an exposed or infectious population larger than 0.011%.

Another important variable to estimate the effect of the incoming exposed and infectious individuals is the effective reproduction number R(*t*) on the day of reopening, column 7 of Table 1, since this number determines how fast the virus will replicate locally within Newfoundland and Labrador. The effective reproduction number on July 1, 2020 was largest in Florida with 2.15, Nevada with 2.10, Montana with 1.88, Idaho with 1.81, and Delaware with 1.80. Of the Canadian provinces and territories, it was only defined in Alberta with 1.31, British Columbia with 1.09, Saskatchewan with 0.44, and Ontario with 0.33. The number of new cases was zero, or too low to calculate a meaningful effective reproduction number, in Newfoundland and Labrador, New Brunswick, Nova Scotia, Prince Edward Island, Manitoba, the Northwest Territories, Nunavut, Quebec, and Yukon. This is why we use the population weighted mean effective reproduction number for Canada, 1.16± 0.17, and for North America, 1.35 ± 0.33 for the reopening forecast in Newfoundland and Labrador.

### Estimated COVID-19 exposed and infectious travelers

Figures 4 and 5 illustrate the daily number of incoming exposed and infectious travelers to the province of Newfoundland and Labrador. The number of exposed and infectious air passengers from Canada and the United States is a reflection of both the frequency of air travel from Figure 2 and the local outbreak dynamics from Figure 3 and Table 1. Figure 4 shows that most exposed travelers come from Florida with 0.137/day, Texas with 0.013/day, Nevada with 0.011/day, Alberta and Ontario both with 0.005/day. Figure 5 shows that most infectious travelers come from Florida with 0.192/day, Texas with 0.023/day, Nevada, Quebec, and Ontario all with 0.016/day. If air traffic would fully resume with the outbreak dynamics of July 1, 2020, the estimated number of incoming exposed and infectious travelers would be 0.203/day and 0.329/day, suggesting that every five days, an exposed traveler, and every three days, an infectious traveler would enter the province of Newfoundland and Labrador via air travel.

**Fig. 4.**
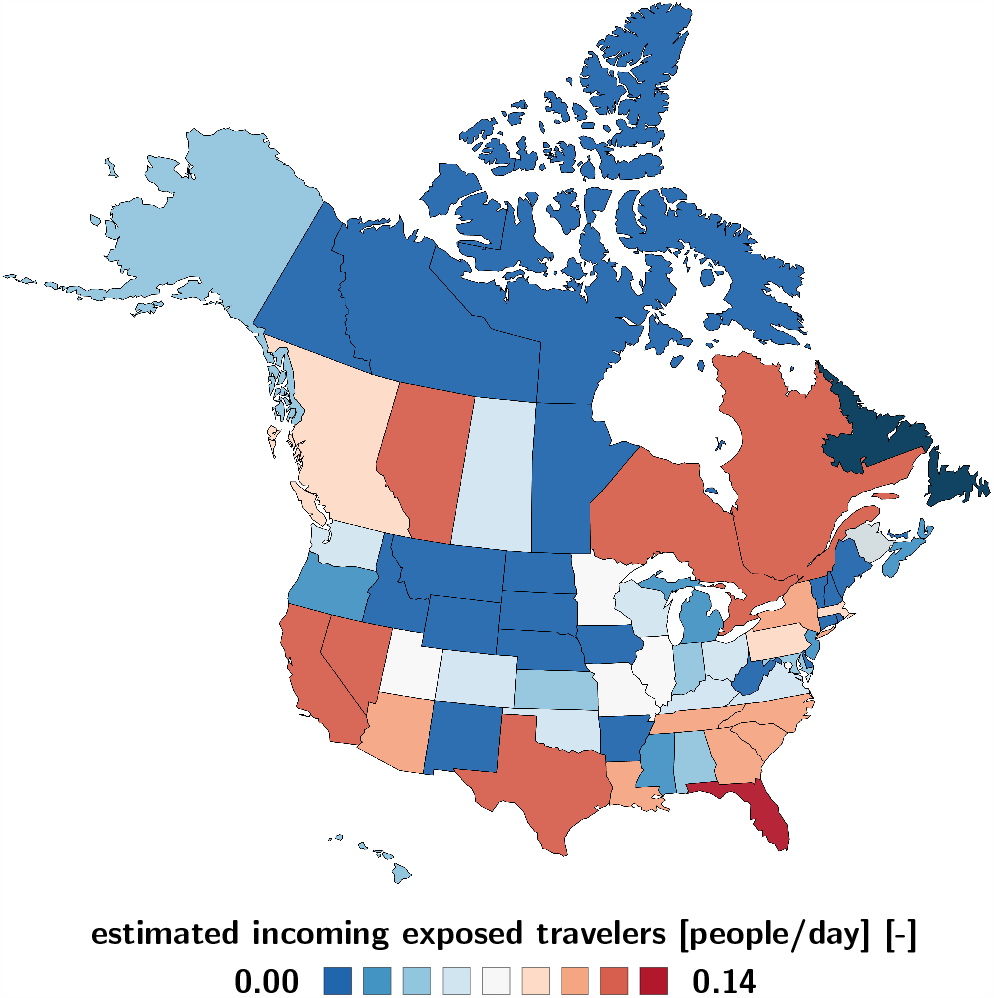
Estimated COVID-19 exposed travelers to Newfoundland and Labrador. Number of daily incoming air passengers from Canada and the United States that have been exposed to COVID-19. The estimate assumes average daily travel from Figure 2 and the outbreak dynamics from Figure 3 as of July 1, 2020.

**Fig. 5.**
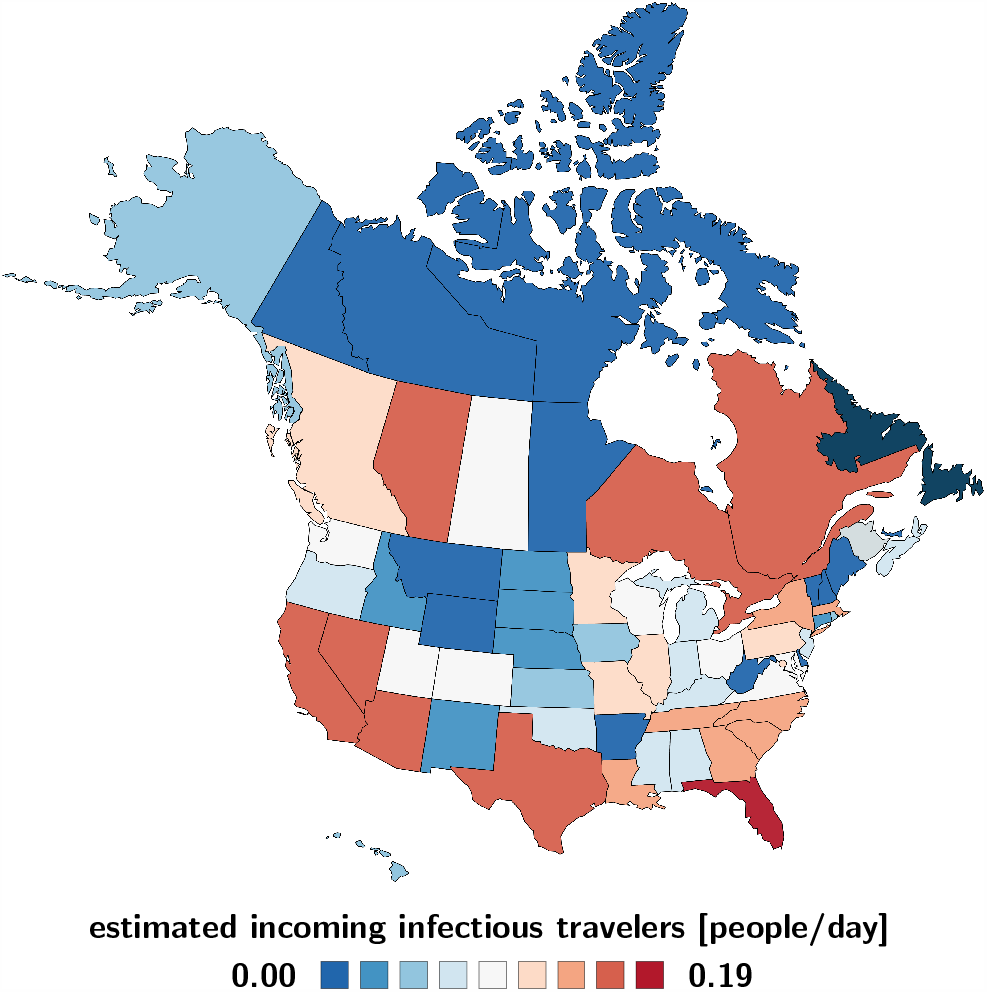
Estimated COVID-19 infectious travelers to Newfoundland and Labrador. Number of daily incoming air passengers from the Canadian provinces and territories and the United States that are infectious with COVID-19 assuming average daily travel from Figure 2 and the outbreak dynamics from Figure 3 as of July 1, 2020.

### Outbreak dynamics of COVID-19 in Newfoundland and Labrador

Figures 6 and 7 highlight the timeline of the COVID-19 outbreak in Newfoundland and Labrador. The graphs distinguish between three time intervals: the first period without travel restrictions from March 15 until May 4, 2020, the second period with travel restrictions from May 4 until July 1, 2020, and the third period upon gradual reopening from July 1, 2020 forward. The graphs report the daily new cases and total cases as dots, compared to the fit of the SEIR model as solid orange curves, from the early outbreak on from March 15 until July 1, 2020. From then on, the dashed lines highlight the reopening forecast for a 60-day period. The solid and dashed lines represent the median values, the shaded areas highlight their 95% confidence intervals.

**Fig. 6.**
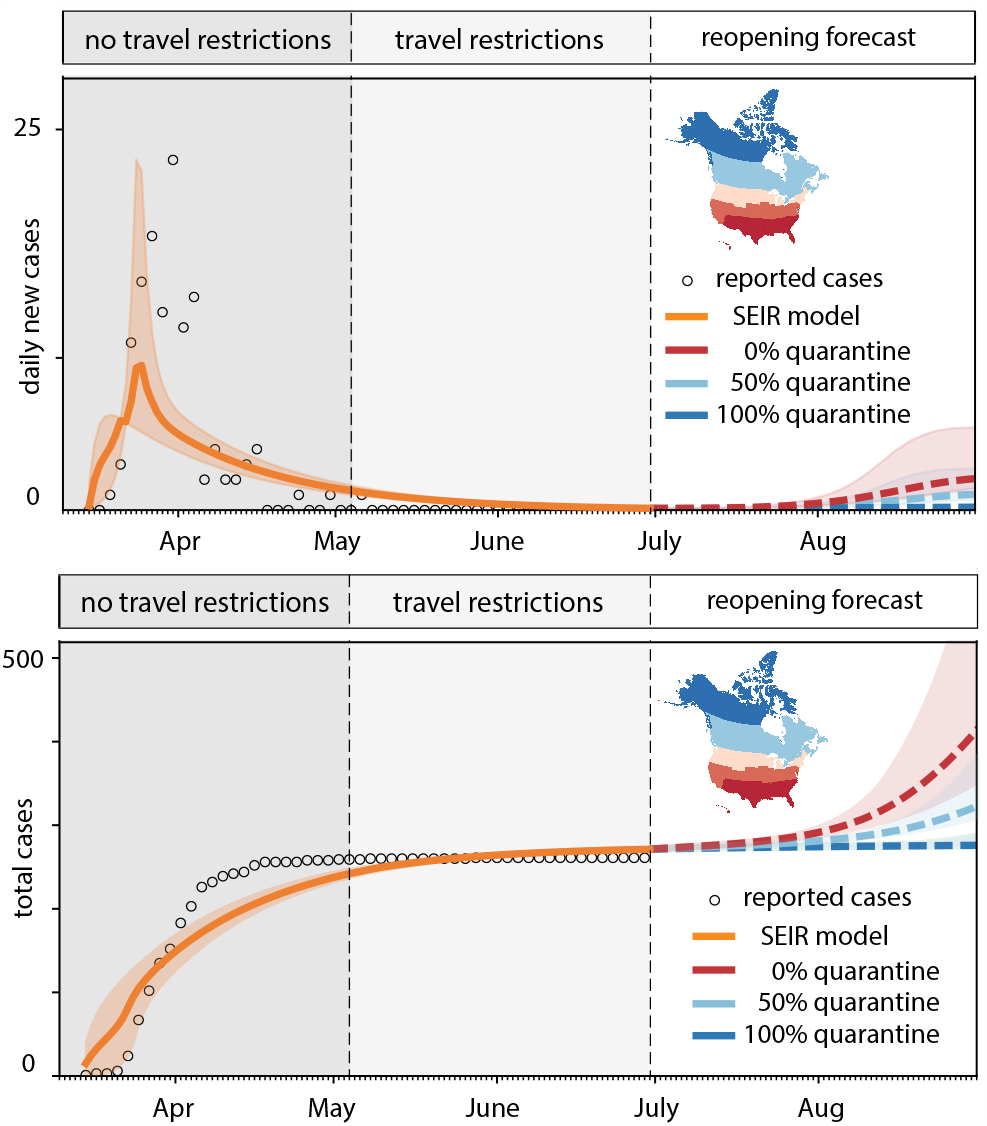
Outbreak dynamics of COVID-19 in Newfoundland and Labrador and the effect of quarantine. Daily new cases and total cases with model fit for periods without travel restrictions and with travel restrictions. Reopening forecast for 60-day period with all incoming travelers quarantining to 100% (dark blue), 50% (light blue), and 0% (red). Solid and dashed lines represent the median values, shaded areas highlight the 95% confidence intervals.

**Fig. 7.**
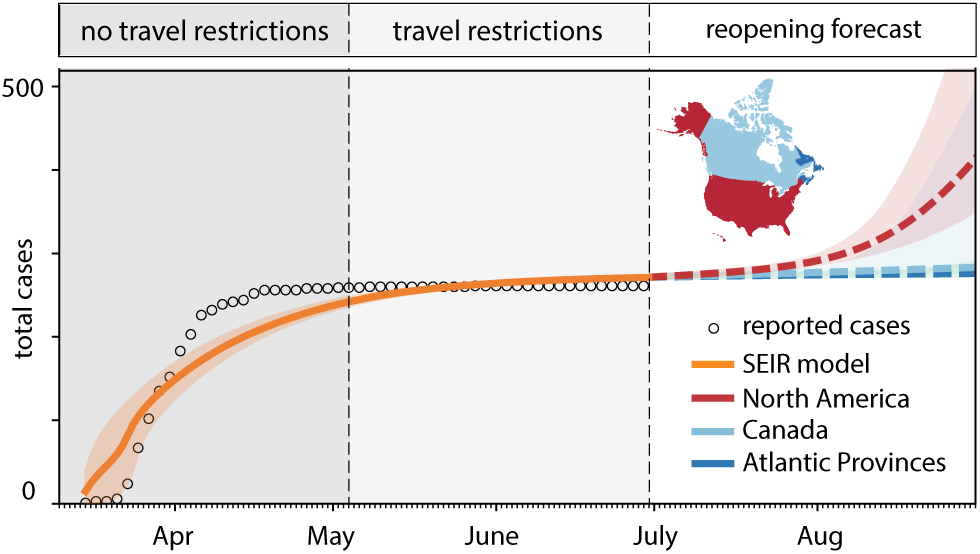
Outbreak dynamics of COVID-19 in Newfoundland and Labrador and the effect of restricted travel. Total cases with model fit for periods without travel restrictions and with travel restrictions. Reopening forecast for 60-day period with mobility only within the Atlantic Provinces (dark blue), all of Canadian (light blue), and all of North America (red). Solid and dashed lines represent the median values, shaded areas highlight the 95% confidence intervals.

Figure 6 explores the effect of quarantine upon reopening: all incoming travelers quarantining to 100% in dark blue, to 50% in light blue, and 0% in red. While a 100% quarantine does not result in any new cases, 50% quarantine results in a mild but notable increase of new cases, and 0% quarantine triggers a rapid exponential outbreak. Figure 7 explores the effect of restricted travel under the assumption of no quarantine: air traffic only between the Atlantic Provinces in dark blue, between all of Canada in light blue, and between all of North America in red. While an opening to the Atlantic Provinces and to all of Canada does not result in a notable number of new cases, under the current conditions, an opening to all of North America would trigger a rapid exponential outbreak.

### Effects of quarantine and restricted travel

Figures 8 and 9 summarize the effects of quarantine and restricted travel on the number of COVID-19 cases in Newfoundland and Labrador for a 150-day period as predicted by our reopening forecast. The solid and dashed lines show the reopening forecast with incoming travelers quarantining at varying percentages and with travel restricted to the Atlantic Provinces, Canada, and all of North America. The predictions use the mean effective reproduction numbers of *R*_t_ = 1.35 for all of North America and *R*_t_ = 1.16 for Canada. The black horizontal lines mark 520 predicted cases corresponding to 0.1% of the population of Newfoundland and Labrador. Table 2 summarizes the dates and number of days by which the curves cross this line and the number of COVID-19 cases reaches 0.1% of the population. Our simulation predicts that this will occur after only 37 or 38 days without travel restrictions and no quarantine. Quarantining only half of the incoming population increases this time period to 44 or 46 days, quarantining 95% of the incoming population to 77 and 89 days. Limiting travel within Canada increases this time period to 95 and 123 days, limiting it within only the Atlantic Provinces to 99 and 128 days.

**Table 2.** Effects of quarantine and restricted travel on COVID-19 dynamics in Newfoundland and Labrador. The dates and days mark the time point at which the number of cases reaches 0.1% of the population. Simulations use the mean effective reproduction numbers of North America and Canada of R_t_ = 1.35 and R_t_ = 1.16 for the prediciton.

| travel from<br>North America | $R_t = 1.35$ | | $R_t = 1.16$ | |
| --- | --- | --- | --- | --- |
|  | date | days | date | days |
| 0% quarantine | 08/07/2020 | 37 | 08/08/2020 | 38 |
| 25% quarantine | 08/10/2020 | 40 | 08/11/2020 | 41 |
| 50% quarantine | 08/14/2020 | 44 | 08/16/2020 | 46 |
| 75% quarantine | 08/22/2020 | 52 | 08/25/2020 | 55 |
| 95% quarantine | 09/16/2020 | 77 | 09/28/2020 | 89 |
| travel at<br>0% quarantine | $R_t = 1.35$ | | $R_t = 1.16$ | |
|  | date | days | date | days |
| North America | 08/07/2020 | 37 | 08/08/2020 | 38 |
| Canada | 10/04/2020 | 95 | 11/01/2020 | 123 |
| Atlantic Provinces | 10/08/2020 | 99 | 11/06/2020 | 128 |

**Fig. 8.**
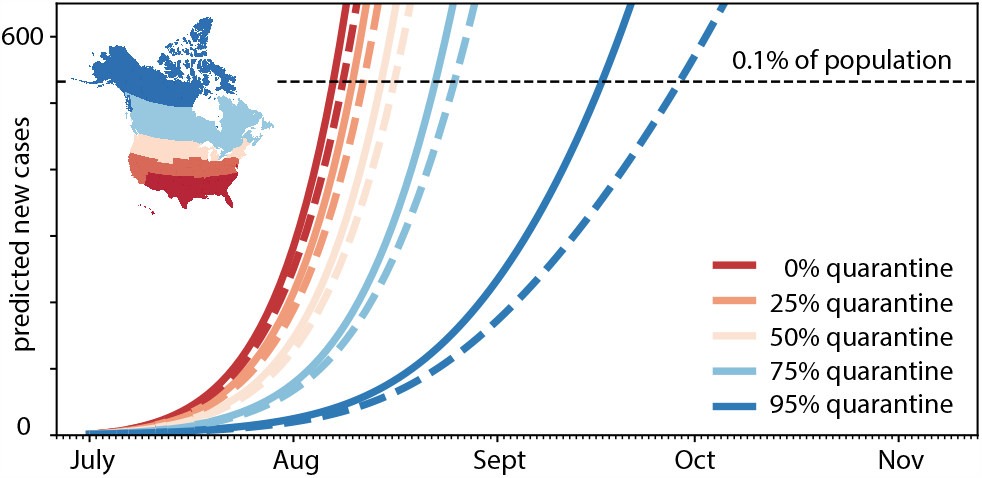
Effect of quarantine on COVID-19 in Newfoundland and Labrador. Reopening forecast for 150-day period with incoming travelers quarantining from 95% (blue) to 0% (red). Predictions are based on a local SEIR model with incoming air traffic from all of North America using the mean effective reproduction number of R_t_ = 1.35 for all of North America (solid lines) and R_t_ = 1.16 for Canada (dashed lines). The black horizontal line marks 0.1% of the population of Newfoundland and Labrador.

**Fig. 9.**
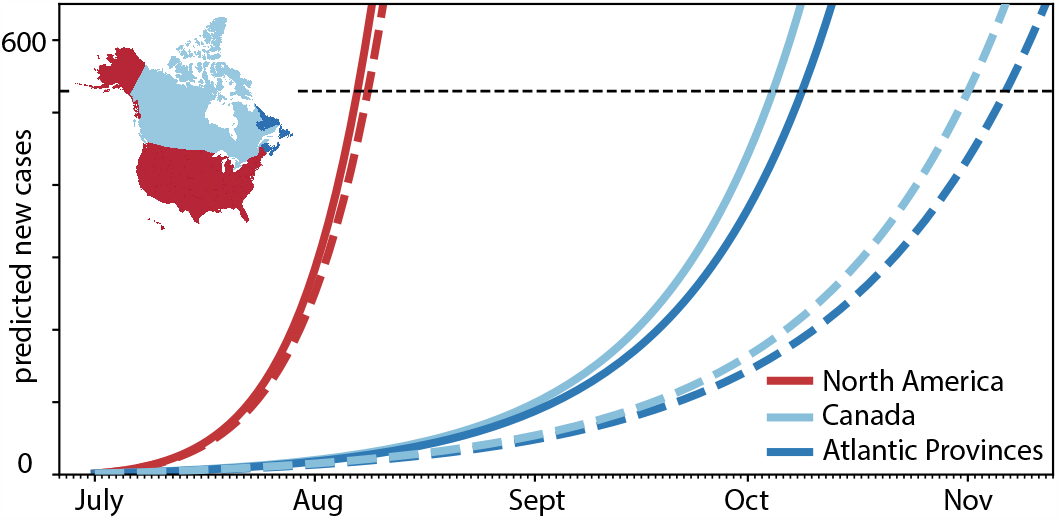
Effect of restricted travel on COVID-19 in Newfoundland and Labrador. Reopening forecast for 150-day period with incoming travelers from Atlantic Provinces, Canada, and all of North America with no quarantine requirements. Predictions are based on a local SEIR model with incoming air traffic from all of North America using the mean effective reproduction numbers of *R*_t_ = 1.35 for all of North America (solid lines) and *R*_t_ = 1.16 for Canada (dashed lines). The black horizontal line marks 0.1% of the population of Newfoundland and Labrador.

## 4 Discussion

Two interacting features determine the outbreak dynamics of the COVID-19 pandemic: the local epidemiology of the disease and the global mobility of diseased individuals [22]. Reducing mobility is a controversial but highly effective measure to manage the outbreak dynamics [40]. Early in the pandemic, many countries have implemented travel restrictions to reduce the number of infectious individuals that travel into the country from the outside [4]. In parallel, they have managed the local epidemiology by reducing contacts through limiting large gatherings, closing schools, or implementing total lockdowns [10]. Some regions, mostly smaller states or provinces, have successfully managed to reduce the number of current cases to zero. One of those provinces is the Canadian province of Newfoundland and Labrador [2].

Obviously, the most effective way to prevent a future outbreak would be to keep isolating these healthy regions from the outside world. The freedom of movement and economic constraints are probably the two strongest arguments for reopening [25]. Numerous different reopening strategies have been proposed, but not all of them seem equally adequate for smaller provinces and states. Here we explore two complementary exit strategies: partial reopening and quarantine. We use the example of Newfoundland and Labrador to forecast the local outbreak dynamics for a region that has not seen any new cases for more than two months.

The disease conditions outside Newfoundland and Labrador change continuously throughout the course of the pandemic and it is critical to dynamically adjust the reopening strategy. Before the travel restrictions were announced on May 4, 2020, the seven-day average of new cases was 1717.0 for Canada with 1091.6 and 438.1 cases originating from the two largest provinces, Quebec and Ontario. By July 1, 2020, these seven-day averages had fallen to 311.1 for Canada, 83.9 for Quebec, and 171.6 for Ontario [6]. In the Atlantic Provinces, the infection rate has remained low throughout the entire period, except for Nova Scotia that had a seven-day average of 12.1 on May 4, 2020 which dropped to 0.3 by July 1, 2020 [6]. Figure 3 provides a side-by-side comparison of these individual case trajectories. Unlike the United States, the Canadian provinces and territories display a clear trend towards slowing the spread of new cases [2]. Notably, all Atlantic Provinces have managed to maintain community transmission at a minimum.

An important piece of information in predicting the effects of partial and total reopening is the daily count of incoming exposed and infectious travelers. We estimate these populations using our network epidemiology model for North America, calibrated with the reported case data for all Canadian provinces and territories [2] and all United States [34], and multiply these numbers with the daily travel data for each region [15]. Table 1 summarizes this information as of July 1, 2020 and Figures 4 and 5 illustrate the estimated number of incoming exposed and infectious travelers per day. These contour plots allow us to quickly identify critical regions like Florida, from which 0.137 exposed and 0.192 infectious individuals would enter Newfoundland and Labrador every day. They also suggest that it is safe to open travel within the Atlantic Provinces, New Brunswick, Nova Scotia, and Prince Edward Island, and reasonably safe to open travel within Canada. While the travel frequency within the Atlantic Provinces and Canada is high as we can see in Figure 2, the current case numbers in these regions are low enough to keep the risk of opening low. The opposite is true for states like California, with a low travel frequency but high case numbers, which suggests that opening travel within all of North America would expose Newfoundland and Labrador to a disproportionally high risk.

An interesting metric is the estimated number of incoming exposed and infectious travelers upon full reopening, 0.203/day and 0.329/day. This suggests that every five days and every three days, an exposed and an infectious traveler would enter the province of Newfoundland and Labrador. In other words every other day, a new COVID-19 case would enter the Newfoundland and Labrador via air travel. Since the exposed and early infectious individuals are still pre-symptomatic, it is impossible to identify and isolate them without strict quarantine requirements [12]. To estimate the precise effects of quarantine and restricted travel, Figures 6 and 7 show our reopening forecast for a 60-day window starting on July 1, 2020. The forecast confirms our intuition that there are two strategies to prevent a new outbreak, either mandating strict quarantine requirements or limited travel within the *Atlantic Bubble*. The increasingly wide 95% confidence intervals for more relaxed conditions suggest that reliable predictions become difficult beyond this time window and case numbers could very well increase beyond control within only a few weeks.

To estimate the time window beyond which the new COVID-19 case numbers would exceed 0.1% of the population, we perform a forward analysis using the data from Table 1 as initial condition and assuming that the exposed and infectious groups maintain their current status quo for all regions except for the province of Newfoundland and Labrador. Figures 8 and 9 show the effects of quarantine and restricted travel for two different effective reproduction numbers that reflect the population-weighted mean across all of North America and across Canada. As such, both Figures showcase the effects of local epidemiology and global mobility: The differences between the solid and dashed curves of the same color are a result of the different reproduction numbers, R_t_ = 1.35 for solid and R_t_ = 1.16 for dashed, associated with new internal cases from within Newfoundland and Labrador [8]; the differences between all solid curves and all dashed curves are a result of varying mobility, associated with new external cases traveling into Newfoundland and Labrador [4]. From the intersection of these curves with the black dashed lines, we can estimate the dates and number of days by which the number of COVID-19 cases reaches 0.1% of the population, which we summarize in Table 2. At full reopening, without travel restrictions and no quarantine, this will occur after only 37 or 38 days. Quarantining only half of the incoming population increases this time period to 44 or 46 days, quarantining 95% of the incoming population to 77 and 89 days. Limiting travel within Canada increases this time period to 95 and 123 days, limiting it within only the Atlantic Provinces to 99 and 128 days. Strikingly, this simple analysis suggests that, under current conditions, banning air travel from outside Canada is more efficient in managing the pandemic than fully reopening and quarantining 95% of the incoming population.

Just like any model, our network epidemiology model has several important limitations: First, the current effective reproduction number in a region with no active cases is undefined which makes predictions very difficult. Here we approximate the effective reproduction number of Newfoundland and Labrador by the population-weighted mean across all of North America and across Canada, which we infer from the reported COVID-19 case data using machine learning [28]. However, we have no idea how well these estimates represent the true behavior of the population of Newfoundland and Labrador upon gradual reopening, since a population that has not experienced a serious outbreak may be more complacent. Second, we assume that the effective reproduction number of all states, provinces, and territories remains constant at its July 1, 2020 level. Since this number reflects both social behavior and personal interaction [7], this is a strong limitation. Yet, it seems reasonable to anticipate that the current North American mean of R_t_ = 1.36 represents an upper limit for our prediction in the sparsely populated province of Newfoundland and Labrador. Third, we have assumed that the fraction of exposed and infectious travelers to Newfoundland and Labrador is similar to the exposed and infected fractions at the locations of origin. Naturally, we would assume that truly sick individuals are less likely to travel, a behavioral effect, which we could include using agent-based modeling. Fourth, our mobility forecast assumes that air traffic fully resumes at its 150-day mean before the outbreak of the pandemic [15]. With people being less likely to travel in the wake of a global pandemic, this estimate might rather represent an upper bound than the true case scenario upon reopening. Fifth, our current model does not include seasonality, which could play a critical role, both in the local disease dynamics and in the global mobility pattern. While we can only estimate the seasonality of the disease, we could certainly include the known economical factors of seasonal fishing and tourism to include the seasonality of mobility. Sixth, our study uses an SEIR model with fixed latency and infectious rates *α* and *γ*. Using a more complex epidemiology model [42] or inferring the rate constants from the reported case data could certainly improve the fit and make the predictions more accurate. Probable even more importantly, the epidemiology model we have used in this study does not include asymptomatic individuals. Asymptomatic transmission plays an important role in spreading COVID-19 [30]. Once seroprevalence data become available for the province of Newfoundland and Labrador, we could include this information and make more accurate predictions about individuals that travel with mild or no symptoms but still carry the virus and could infect others.

## 5 Conclusion

Relaxing travel restriction is a highly contentious political decision. However, from an outbreak dynamics perspective, the picture is quite clear: Without proper control, an influx of infected travelers can easily become the seed for a new exponential outbreak. In the early phases of exponential growth, the new case numbers may appear low and manageable, but when unaddressed, the case numbers will begin to grow at an alarming rate. At this point, it becomes impossible to manage a new outbreak with soft measures alone. Clearly, opening borders is a critical decision, especially towards regions with a higher case prevalence. Our study shows that– especially for smaller provinces or states–tight border control is often easier and more effective than quarantine. Partial reopening, for example within local travel bubbles, is an effective compromise and a reasonable first step. Our results suggest that relaxing travel restrictions entirely is possible, but would require strict quarantine conditions. Voluntary quarantine, even at an overall rate of 95%, is not enough to entirely prevent future outbreaks. A solution to reduce the quarantine time is to test, trace, and isolate. Without these policies, even regions that have successfully managed the COVID-19 pandemic to date are at risk of seeing a new epidemic outbreak within only a few weeks. This outbreak can quickly grow out of control and require extensive and expensive lockdown periods and other stringent non-pharmaceutical interventions. It is therefore important to understand, closely monitor, and predict the local and global outbreak dynamics and be aware of the dangers associated with uncontrolled reopening.

## Data Availability

all data are available on public data bases cited in the text

## Acknowledgements

We acknowledge stimulating discussions with Dr. Mathias Peirlinck and Dr. Francisco Sahli Costabal. This work was supported by a DAAD Fellowship to Kevin Linka, the Engineering and Physical Sciences Research Council grant EP/R020205/1 to Alain Goriely, and a Stanford Bio-X IIP seed grant and the National Institutes of Health Grant U01 HL119578 to Ellen Kuhl.

